# Safety and immunopotency of an adenovirus-vectored tuberculosis vaccine delivered via inhaled aerosol to healthy humans: a dose and route comparison phase 1b study

**DOI:** 10.1101/2021.09.09.21263339

**Authors:** Mangalakumari Jeyanathan, Dominik K. Fritz, Sam Afkhami, Emilio Aguirre, Karen J. Howie, Anna Zganiacz, Anna Dvorkin-Gheva, Michael R. Thompson, Richard F. Silver, Ruth P. Cusack, Brian D. Lichty, Paul M. O’Byrne, Martin Kolb, Maria Fe C. Medina, Myrna B. Dolovich, Imran Satia, Gail M. Gauvreau, Zhou Xing, Fiona Smaill

**Affiliations:** McMaster Immunology Research Centre, McMaster University, Hamilton, Ontario, Canada; M. G. DeGroote Institute for Infectious Disease Research, McMaster University, Hamilton, Ontario, Canada; Department of Medicine, McMaster University, Hamilton, Ontario, Canada; Department of Pathology & Molecular Medicine, McMaster University, Hamilton, Ontario, Canada; Department of Chemical Engineering, McMaster University, Hamilton, Ontario, Canada; Department of Critical Care and Sleep Medicine, Case Western Reserve University School of Medicine, Cleveland, Ohio, USA

**Author notes:** **Joint lead & correspondence contacts:** (Z. Xing), (F. Smaill).

## Abstract

**Background:** Adenoviral (Ad)-vectored vaccines are typically administered via intramuscular injection to humans, but this route of delivery is unable to induce respiratory mucosal immunity which requires respiratory mucosal route of vaccination. However, inhaled aerosol delivery of Ad-vectored vaccines has remained poorly characterized and its ability to induce respiratory mucosal immunity in humans is still unknown. The goal of our study was to evaluate and compare the safety and immunogenicity of a human serotype 5 Ad-based tuberculosis (TB) vaccine (AdHu5Ag85A) delivered to healthy humans via inhaled aerosol or intramuscular injection.

**Methods:** In this open-labeled phase 1b trial, 31 healthy adults between 18 and 55 years of age with a history of BCG vaccination were enrolled at McMaster University Medical Centre, Hamilton, Ontario, Canada. AdHu5Ag85A was administered by a single-dose aerosol using the Aeroneb® Solo Vibrating Mesh Nebulizer or by intramuscular (IM) injection; 11 in the low dose (LD, 1×10^6^ PFU) aerosol group, 11 in the high dose (HD, 2×10^6^ PFU) aerosol group and 9 in the IM (1×10^8^ PFU) group. The primary outcome was safety of a single administration of vaccine delivered to the respiratory tract by aerosol or by IM injection. The vaccine-related local and systemic adverse events were collected from participants from a self-completed diary for 14 days after vaccination and at scheduled follow-up visits. Routine laboratory biochemical and haematological tests were measured at 2, 4 and 12 weeks after vaccination and lung function was measured at 2, 4, 8 and 12 weeks after vaccination. The secondary outcome was comparison of immunogenicity among the different routes and aerosol dose groups. Immunogenicity to aerosol or IM vaccination was measured both in the peripheral blood and bronchoalveolar lavage samples by Luminex, and cell surface and intracellular cytokine immunostaining. Anti-AdHu5 antibodies and neutralization titers were determined before and after vaccination using ELISA and bioassay, respectively. This trial is registered with ClinicalTrial.gov, NCT02337270.

**Results:** The aerosol droplets generated by Aeroneb® Solo Nebulizer were mostly <5.39µm in size, suitable for efficient Ad-vectored vaccine deposition to major human airways. Both LD and HD of AdHu5Ag85A administered by aerosol inhalation and the intramuscular injection were safe and well-tolerated. Respiratory adverse events were infrequent, mild, transient and similar among groups. IM injection was associated with a mild local injection site reaction in two participants. Systemic adverse events were also infrequent, mild, transient and similar among all groups. There were no grade 3 or 4 adverse events reported nor any serious adverse events. Both aerosol doses, particularly LD, but not IM, vaccination markedly induced Ag85A-specific airway tissue-resident memory CD4 and CD8 T cells of polyfunctionality. While as expected, IM vaccination induced Ag85A-specific T cell responses in the blood, the LD aerosol vaccination also elicited such T cells in the blood. Furthermore, the LD aerosol vaccination induced persisting transcriptional changes in alveolar macrophages indicative of trained innate immunity.

**Interpretation:** Inhaled aerosol delivery of Ad-vectored vaccine is a safe, economical and superior way to elicit respiratory mucosal immunity. The results of this study encourage further development of aerosol vaccine strategies against not only TB but also other respiratory pathogens including COVID-19.

**Funding:** The Canadian Institutes for Health Research and the Natural Sciences and Engineering Research Council of Canada.

**Research in context:** *Evidence before this study:* We searched PubMed for published research articles without language or date restrictions by using search terms “adenovirus”, “aerosol” and “clinical trial” or “virus”, “vaccine”, “aerosol”, and “clinical trial”. Our search results indicate that adenoviral-vectored vaccine has rarely been delivered via inhaled aerosol into the respiratory tract of healthy humans. Adenoviral-vectored TB or HIV vaccine was delivered via inhaled aerosol to non-human primates, providing support for its further development for human application. An aerosolized adenoviral vector expressing CFTR was tested as a gene replacement therapeutic in cystic fibrosis patients. Recently, although an adenoviral-vectored COVID-19 vaccine was delivered via aerosol to humans, there were no evidence presented that it induced local mucosal immunity. Given the increased recognition of its value, inhaled aerosol has been explored in humans with non-adenoviral, viral vaccines against respiratory infections of global importance including measles and TB. However, since different aerosol technologies were used in these studies and aerosol characteristics including delivery efficiency vary according to the viral platform, aerosol delivery technology and its efficiency in inducing respiratory mucosal immunity remain to be established for administering adenoviral-vectored vaccine to healthy humans.

*Added value of this study:* This represents the first study to demonstrate the characteristics of a single-dose aerosolized human serotype 5 adenoviral-vectored vaccine, and its safety and immunogenicity in BCG-vaccinated healthy humans. It is also the first to show the superiority of inhaled aerosol immunization with this type of vaccine platform, over intramuscular (IM) route of immunization, in inducing respiratory mucosal immunity. Robust adaptive immune memory responses were induced in the respiratory tract with an aerosol vaccine dose up to 100 times smaller than the dose for IM immunization. Besides local mucosal immunity induced by aerosol immunization, it also induces levels of systemic immunity comparable to those by IM immunization. Also for the first time, we show that contrast to IM immunization, aerosol immunization does not increase either local or systemic anti-adenoviral neutralizing antibodies.

*Implications of all the available evidence:* Collectively our findings show the technical feasibility, safety and potency of needle/pain-free delivery of a recombinant adenoviral-vectored vaccine to the respiratory tract of healthy adults. This vaccine strategy differs from parenteral route of immunization in its potency to elicit much desired respiratory mucosal immunity consisting of trained macrophages and tissue-resident memory T cells. The study provides the proof of concept to endorse inhaled aerosol vaccine strategies against pulmonary TB. Of further importance, as a number of currently authorized COVID-19 vaccines are also adenoviral-vectored, our study offers important technological details and scientific rationale for the development of inhalable next-generation COVID-19 vaccines aiming to induce all-around protective respiratory mucosal immunity in humans.

## Introduction

Pulmonary tuberculosis (TB) continues to be a major global health issue, accounting for 1.4M deaths and 10M new cases in 2019 ^1^. BCG, as the most administered human vaccine which is given via the skin shortly after birth, has failed to effectively control TB in adults. A safe and effective boost vaccine strategy is urgently needed for much improved protective immunity in the lung ^2, 3^.

Among the promising vaccine platforms is a recombinant replication-defective human serotype 5 adenoviral-vectored TB vaccine expressing *M.tb* antigen 85A (AdHu5Ag85A). This vaccine has been extensively evaluated in a number of preclinical models, shown to be highly effective when administered via the respiratory tract, as opposed to its parenteral delivery ^3, 4^. Besides its superior effects in inducing lung tissue-resident memory T cells ^3^, respiratory mucosal-delivered AdHu5Ag85A is able to elicit long-lasting memory airway macrophages and trained innate immunity ^5, 6^. It is widely believed that the most effective vaccine strategy ought to induce both innate memory and adaptive memory responses ^5, 7^. Although AdHu5Ag85A was evaluated successfully in healthy humans following intramuscular injection^8, 9^, its suitability for respiratory mucosal delivery and its safety and immunogenicity remain to be determined in BCG-vaccinated healthy humans.

Recent studies have shown inhaled aerosol to be a safe and effective delivery method for respiratory mucosal route of immunization in healthy humans with measles and MVA85A vaccines ^10–13^. However, these studies applied different technologies for aerosol delivery. Since aerosol characteristics and delivery efficiency may vary according to the type of vaccine, an aerosol delivery technology remains to be characterized and validated for administering adenoviral-vectored vaccine to the human airway. Although an adenoviral-vectored COVID-19 vaccine was delivered via aerosol to humans in a recent clinical trial ^14^, unfortunately its ability and advantage to induce respiratory mucosal immunity was not assessed and thus remains unknown. Given that a number of currently approved COVID-19 vaccines are also based on adenoviral vector, it is highly relevant to fully characterize an inhaled aerosol delivery technology for adenoviral-vectored vaccine and investigate its ability to induce respiratory mucosal immunity in preparation for its translation to respiratory mucosal COVID-19 vaccine strategies ^15^. Such next-generation COVID-19 vaccine strategies are urgently needed in the face of increasing break-through infections due to the variants of concern and waning vaccine-induced immunity ^16^.

In the present phase 1b study, we have characterized the property of AdHu5Ag85A aerosol droplets generated by the AeroNeb Solo nebulizer. We evaluated the two aerosol doses and compared the safety and immunogenicity of the vaccine delivered via the respiratory mucosal route or intramuscular route in previously BCG-vaccinated healthy adults. Our study is the first to safely deliver an adenovirus-vectored vaccine via inhaled aerosol to humans and to demonstrate its superiority in inducing respiratory mucosal immunity over intramuscular injection.

## Methods

### Study design and participants

This was an open-labeled phase 1 trial investigating a single recombinant genetic TB vaccine AdHu5Ag85A in healthy subjects with a history of BCG vaccination. The vaccine was administered as a single dose either by inhaled aerosol or by intramuscular (IM) injection. Study participants were recruited by advertising, approved by the local REB. BCG (+) status was determined based on patient/parent recall, vaccination records, or birth country where BCG vaccine is routinely administered (Fig 1). All enrolled participants were healthy and had normal baseline hematology, spirometry and DLCO, biochemistry, chest X-ray and negative serological testing for HIV antibody. Latent *Mtb* infection was excluded by a negative interferon gamma release assay. Current smokers and ex-smokers who had quit within the last year, people with a history of inhaled recreational drugs, respiratory disease, e.g., asthma, chronic obstructive pulmonary disease (COPD), and pregnant or lactating women were excluded. Following vaccination, participants were asked to record their temperature twice a day daily at set times for 5 days and kept a diary to record any symptoms they experience for 14 days. Safety and medical evaluation was performed at baseline, and 48-72 hours, 2, 4, 8, 12, 16 and 24 weeks after vaccine administration. Adverse events were assessed according to the CTCAE Expanded Common Toxicity Criteria. Immunological evaluation was performed with bronchoalveolar lavage and blood samples. Bronchoscopy was carried out within 1 to 6 days before planned vaccination and at both 2 and 8 weeks post-vaccination. Blood samples were collected at baseline and at 2, 4, 8 and 12 weeks post-vaccination.

**Figure 1.**
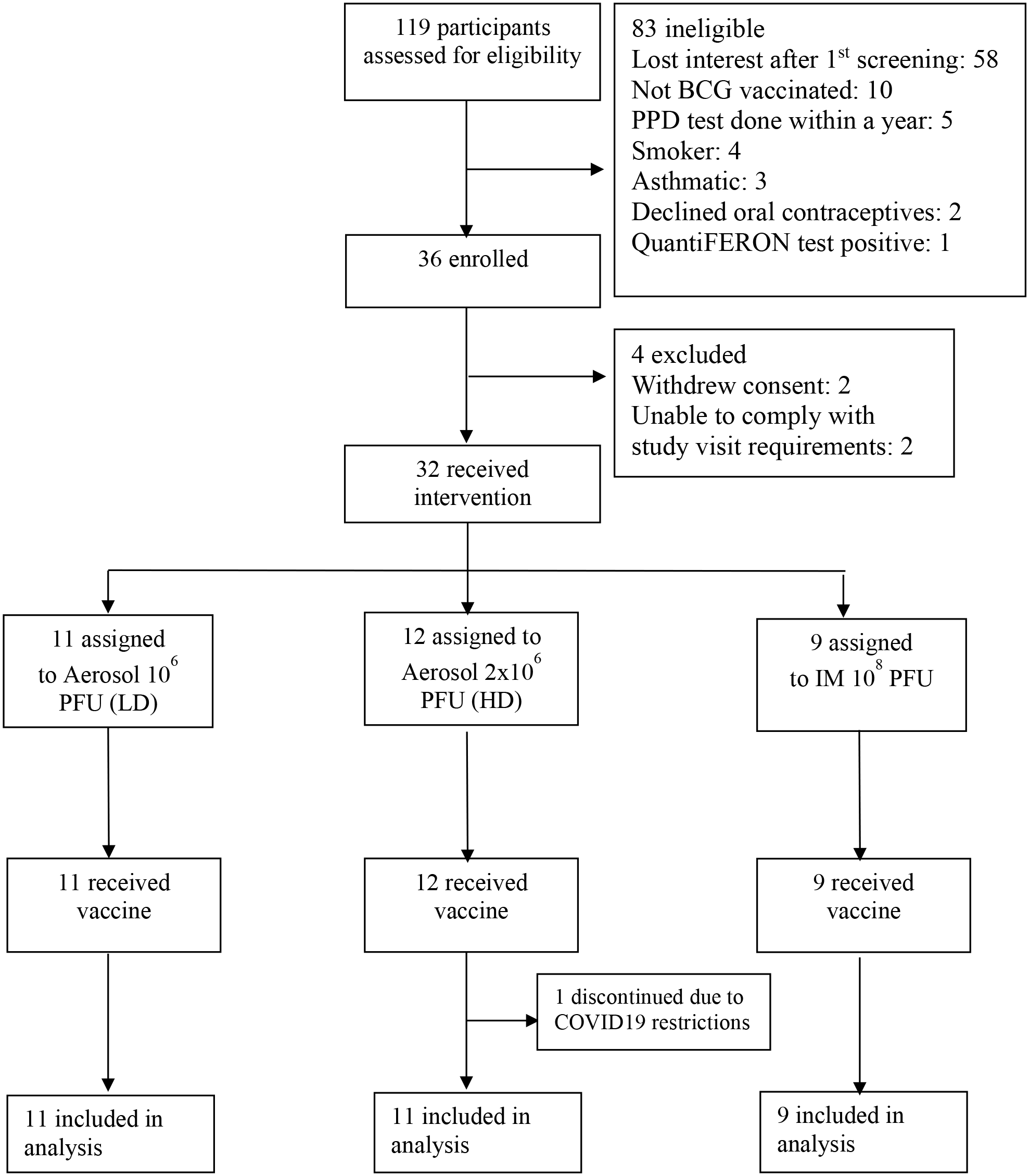
Trial profile. PPD: purified protein derivative; LD: low dose aerosol; HD: high dose aerosol; IM-intramuscular injection; PFU: plaque-forming unit

All participants provided written informed consent. This phase 1 trial was approved by the Health Canada and Hamilton Integrated Research Ethics Board. This trial is registered with ClinicalTrial.gov, number NCT# 02337270.

### Randomization and masking

The first cohort of 8 BCG positive subjects received 10^6^ pfu (vp/pfu ratio: 32)(low dose-LD) AdHu5Ag85 vaccine administered by aerosol. The second cohort consisted of 18 participants randomly allocated to receive either 2×10^6^ pfu (high dose-HD) by aerosol (n=9) or single intramuscular administration of 10^8^ pfu (IM) AdHu5Ag85 vaccine (n=9). For the second cohort, a randomization list was generated containing sequential codes linked to route of study vaccine assignment, either IM injection or aerosol (Fig 1). Following a review of the data after the enrolment of the second cohort, the protocol was amended, and an additional 3 participants were enrolled in the high dose aerosol group and an additional 3 participants were enrolled in the low dose group. The study was not blinded. Each participant served as their own control (before and after vaccination) and there was no placebo group. For safety reasons, 2 participants were first vaccinated for each aerosol dose, and followed for 2 weeks after vaccine administration before immunizing the rest of the participants in the dose group. Reports detailing AEs and SAEs for 4 weeks post-dose were reviewed by the safety monitoring committee before moving to the higher dose. All participants were followed for a total of 24 weeks after vaccine administration.

### Procedures

The Aeroneb® Solo Micropump was selected as the delivery device (Fig S1). The aerosol performance of the Aeroneb® Solo Vibrating Mesh Nebulizer with AdHu5Ag85A was characterized using standard procedures and performance metrics ^17^. Fill volume (FV), delivery time, emitted dose (ED) of vaccine aerosol available at the mouth collected on standard filters, aerosol droplet size characteristics using the NGI Cascade Impactor operated at 15Lpm were measured. Salbutamol sulphate served as the tracer for saline droplets containing vaccine particles. Regional deposition of vaccine droplets in the lung was estimated from the particle size metrics of the carrier (salbutamol) aerosol. An indication of the available vaccine dose at the mouth was predicted from ED. The amount of aerosol-containing vaccine estimated to be deposited into the lung was calculated using ED in combination with particle size statistics. Viability of the aerosolized vaccine was determined by plaque forming assay.

Clinical grade AdHu5Ag85A vaccine was provided by the Robert E Fitzhenry Vector Laboratory, McMaster Immunology Research Centre, McMaster University, Hamilton, Ontario, Canada. AdHu5Ag85A was produced according to current Good Manufacturing Practices (cGMP) in the Vector Laboratory and has been fully certified. For each participant allocated to receive vaccine by aerosol, a single dose of AdHu5Ag85A diluted in 0.5 ml saline was aerosolized using the Aeroneb Solo and inhaled via mouthpiece using tidal breathing. For participants allocated to receive IM route, AdHu5Ag85A diluted in 0.5 ml saline was injected as described previously ^8^.

Bronchoscopy was performed using a flexible bronchoscope, with the procedure performed in the research facility at the Health Science Centre, McMaster University by a trained respiratory physician. Following light sedation (using midazolam and fentanyl) and local anaesthesia to the upper and lower respiratory tract, the bronchoscope was advanced until wedged in the right middle bronchus and approximately 40ml of sterile saline instilled and then aspirated back using gentle manual suction in a 50 ml syringe. This was sequentially repeated an additional 3 times with a total of 160 ml of saline lavage. Oxygen saturations were monitored throughout the whole procedure. After the bronchoscopy, vital signs (SaO2, HR, BP) were measured immediately, 15 mins, 30 mins, 45 mins, 60 mins and 2 hours during recovery. Spirometry was repeated to ensure FEV1>70% predicted and within 15% of pre-bronchoscopy values before discharge. Each aspirate was kept separate on ice and processed within an hour after collection at McMaster Immunology Research Center. The first aspirate was discarded after obtaining the cell count. 2^nd^, 3^rd^ and 4^th^ BALF was saved separately and stored at −80°C for future analysis. Cells were then pooled and counted.

Interferon gamma release assay was performed using QuantiFERON TB Gold in tube or QuantiFERON TB Gold Plus (Qiagen) according to manufacturer’s instruction to determine latent TB status ^8^. Collected serum and concentrated BALF samples (20-fold concentrated from 10 ml of combined aspirates 2 and 3 using Centriprep 3kDa Cut-off centrifugal filter unit-Millipore-Sigma) were measured for anti-AdHu5 IgG antibodies as previously described ^8^. AdHu5-neutralizing antibody (nAb) levels in serum and concentrated BALF were assessed as a function of GFP-expressing AdHu5 infection using A549 cells as previously described ^18^. For both total IgG and nAb titers, serum samples at a 1:10 and BALF at a 1:5 dilution were used.

We counted the total cells in the BALF and calculated the number of cells per ml of BALF. Differential cell count was performed on the cells from BALF using cytospin. We quantified the Ag85A- and *M.tb* antigen-specific T cell responses in the peripheral blood and in the airways represented by the BALF using ELISA and/or intracellular cytokine staining (ICS) assay as previously described ^8^. Antigens used for stimulation included the pooled Ag85A overlapping peptides (Ag85A p. pool) and a cocktail of mycobacterial antigens consisting of *M.tb* culture filtrate proteins (*M.tb*CF) and recombinant Ag85A protein (rAg85A) at concentrations previously described ^8^. Unstimulated and PHA-stimulated cells were set up in parallel as background and positive controls. For whole blood culture and the ELISA, 1 ml of heparinized whole blood was added into each well of a 24-well plate. Each was stimulated for 18 to 24 hours with one of the antigens mentioned above. Collected plasma was stored at −70°C. Cytokines were determined for human IFNγ, IL-2, and TNFα using Luminex Multiplex Kit from Millipore. Fresh PBMC and BALF cells were stimulated with antigens and wells were processed according to manufacturer’s instruction (BD Biosciences).

For determination of CD4 and CD8 T cell responses to vaccine using the whole blood culture ^10^, 1ml of blood was stimulated with each of the antigens mentioned above in the presence of 1 μg/mL αCD28, 1 μg/mL αCD49d (BD Biosciences). Samples were incubated at 37°C in 5% CO_2_ for 6h with Ag85A p. pool or PHA stimulation or for 12h with *Mtb*CF-rAg85A stimulation. Brefeldin-A was added for last 5h (Ag85A p. pool or PHA) or last 6h (*Mtb*CF-rAg85A or unstimulated whole blood). At the end of incubation, red blood cells were lysed, and samples were frozen in liquid nitrogen until FACS analysis. To determine the CD4 and CD8 T cell responses in the airways, 0.5-1×10^6^ BALF cells were plated and stimulated as for the whole blood. At the end of incubation, cells were immediately stained for ICS and FACS analysis.

Surface immunostaining and ICS was done as previously described ^8^. Briefly, frozen cells from whole blood were thawed and permeabilized before staining with a cocktail of fluorochrome-conjugated monoclonal antibodies; CD3 (FITC), CD4 (PB), CD8 (PECy7), IFNγ (PE), IL-2 (APC), and TNFα (PerCP-Cy5.5). BALF cells were surface-immunostained for viability (Molecular Probes LIVE/DEAD fixable stain (Aqua), Invitrogen) followed by CD4 (AF700), CD14 (V450), CD19 (V450). Cells were then permeabilized and stained for intracellular cytokines IFNγ (PE), IL-2 (APC), and TNFα (FITC). Ag85A p. pool-stimulated whole blood and BALF cells were also stained for surface markers CD103 (APC), CD69 (PerCP-Cy5.5) and CD49d (PE-dazzle) to evaluate the surface expression of tissue-resident memory T cell- or T cell lung trafficking-associated molecules. Cells were analyzed with LSRII flow cytometer and assessed with Flowjo version 9.9.6 (Tree Star Inc.).

We evaluated the induction of trained innate immunity in airway macrophages in the LD dose group. After reviving the frozen BALF cells for 6h, one million viable cells were seeded on a 12-well plate and incubated for 2h at 37^◦^C, 5% CO2. At this point, airway macrophages were enriched by removal of non-adherent cells via extensive washing with pre-warmed (37^◦^C) PBS and then cultured with or without *M.tb* lysates stimulant for 12h. Total RNA was extracted using RNeasy Plus Mini Kit from Qiagen which includes gDNA eliminator columns, following the manufacturer’s protocol. Quality of RNA was verified, and subsequent RNA sequencing was carried out by Farncombe Metagenomic Facility at McMaster University. RNA integrity was checked using the Agilent bioanalyzer. mRNA was converted to cDNA after enrichment. cDNA libraries were sequenced using an Illumina HiSeq machine (1×50 bp sequence reads). Data were analyzed as previously described ^6^. Before the analysis of differentially expressed genes (DEGs), reads were filtered by quality and counted. Genes, showing low levels of expression were removed using EdgeR package in R. Statistical analysis was performed with 11,848 genes. The Limma package in R was used to identify DEGs in stimulated macrophages and DEGs were then compared to their respective unstimulated controls. Stringent criteria, including Log_2_ of fold-change ≥1.5 or ≤1.5, and corrected p value <0.05 were applied to filter DEGs. Ontology analysis for the Biological Processes component was performed using BINGO plugin, and barcharts were created using top 10 biological processes shared between the comparisons (reflecting adjusted p-values averaged between the pairwise comparisons) or using all biological processes unique to the comparison of interest.

### Outcomes

The primary outcome of this trial was safety of a single administration of vaccine delivered to the respiratory tract by aerosol or intramuscular injection. The frequency and severity of vaccine-related local and systemic adverse events were collected from participants from a self-completed diary for 14 days after vaccination (fever, chills, cough, wheezing, sneezing, shortness of breath, chest pain, headache, fatigue or malaise, conjunctivitis, rhinitis, epistaxis, injection site reaction, syncope or light-headedness) and at scheduled follow-up visits.

Routine laboratory biochemical and haematological tests (CBC, sodium, potassium, creatinine, AST, bilirubin) were measured at 2, 4 and 12 weeks after vaccination and lung function (FEV1 and FVC) were measured at 2, 4, 8 and 12 weeks after vaccination. The secondary outcome was comparison of immunogenicity among the different routes and aerosol dose groups.

### Statistical analysis

Adverse events were reported descriptively. Immunogenicity data were analyzed using Prism (version 9·2.0). Data are expressed as the mean value (horizontal line) with 95% confidence interval. Box plots show mean value (horizontal line) with 95% confidence interval (whiskers) and boxes extend from the 25^th^ to 75^th^ percentiles. Violin plots show the median and quartiles. Pie chart shows median proportions of polyfunctional Ag-specific T cells. Wilcoxon matched pairs signed rank test was used when comparing change of T cell or cytokine responses at various timepoints from the baseline values within the same vaccination group (LD aerosol, HD aerosol and IM groups) Mann-Whitney U test was used for comparison of the difference in T cell responses between vaccination groups. For correlation analysis, the Spearman rank coefficient test was used. Two-tailed p values of less than 0.05 were considered significant.

## Results

During the period of March 2019 to February 2021, we enrolled 36 BCG-vaccinated healthy adults between 18 and 55 years of age at McMaster University Medical Centre, Hamilton, Ontario, Canada. 4 participants were excluded (two withdrew consent and two were withdrawn before vaccination because they were unable to comply with the study visit requirements) and one did not complete any follow-up visits after vaccination because of COVID restrictions. 31 participants completed the study, 11 in the low dose aerosol group, 11 in the high dose aerosol group and 9 in the intramuscular group (Fig 1). The demographic and baseline characteristics of the study participants were similar among study groups (Table 1).

**Table 1.** Demographics of participants

|  | **Low dose**<br>**Aerosol 1x10<sup>6</sup>**<br>**N=11** | **High dose**<br>**Aerosol 2x10<sup>6</sup>**<br>**N=12** | **IM 1x10<sup>8</sup>**<br>**N= 9** |
| --- | --- | --- | --- |
| Male | 2 (18%) | 3 (25%) | 7(78%) |
| Female | 9 (82%) | 9 (75%) | 2 (22%) |
| Years (age) | 26 (21-37) | 26 (18-38) | 27 (18-42) |
| BMI (Kg/m2) | 24.1 (17.2-35.1) | 21.9 (18-30.9) | 24.9 (20.1-31.3) |
| BCG vaccination history |  |  |  |
| Participant Recall | 7 (64%) | 5 (41.6%) | 3 (33.3%) |
| Vaccination Record | 1 (9%) | 3 (25%) | 1 (11%) |
| Continent of Birth |  |  |  |
| Asia | 6 (54.5%) | 5 (41.6%) | 4 (44.4%) |
| Europe | 1 (9%) | 1 (8.3%) | 2 (22.2%) |
| America | 1 (9%) | 2 (16.6%) | 3 (33.3%) |
| Middle East | 2 (18.2%) | 2 (16.6%) | 0 |
| India | 1 (9%) | 2 (16.6%) | 0 |
| Spirometry |  |  |  |
| % predicted FEV1 | 100% (96-126) | 99% (89-114) | 106% (81-129) |
| % predicted FVC | 98% (82-130) | 101% (93-122) | 108% (90-130) |
| Median baseline SpO2 | 99% (97-100) | 99% (98-100) | 98% (97-100) |

### Characterization of inhaled aerosol delivery method and aerosol droplets using Aeroneb® Solo device

The Aeroneb® Solo Micropump was selected to be part of the device set up for aerosol generation and delivery in our study (Fig S1). A fill volume of 0.5 ml in the nebulizer was determined to be optimal for vaccine delivery in saline. Subjects completed inhalation of this volume containing the vaccine via tidal breathing in approximately 2.5 minutes (Table 2). The emitted dose of vaccine available at the mouth was found to be approximately 50% of the loaded dose in the nebulizer (Table 2). The majority of aerosol droplets were <5.39 µm (85%) or between 2.08 and 5.39 µm in diameter conducive to vaccine deposition in major airways. Thus, the amount of aerosol available at the mouth and subsequently deposited in the lung was 42.5% ^17^. The estimated rate of viable vaccine from aerosol droplets generated by the nebulizer was 17.4%. The dose loaded in the nebulizer for aerosol inhalation was thus corrected according to the estimated losses of vaccine within the device.

**Table 2.** Characterization of aerosol device and aerosol droplets

| Parameters measured |  |
| --- | --- |
| Nebulizer fill volume | 0.5 ml saline |
| Time to complete nebulization via tidal breathing<br>N=8 (4 male & 4 female adults) | 149.25 ± 8.0 (seconds) |
| Emitted dose at mouth<br>determined with salbutamol sulphate with vaccine using breath simulator<br>(Vt 500 ml, 15bpm)<br>N=4 | 48.8 ± 2.22% |
| Aerodynamic droplet size<br>determined with salbutamol sulphate with vaccine by using the Next<br>Generation Impactor (NGI)<br>N=4 | <5.39 μm: 85.2 ± 2.04%;<br>2.08-5.39 μm: 68.0 ± 1.99% |
| Vaccine viability<br>determined by viral plaque forming unit assay<br>(average value of two experiments) | 17.4% |

### Safety of inhaled aerosol and intramuscular injected AdHu5-Ag85A vaccine

Both low and high doses of AdHu5Ag85A administered by aerosol inhalation or the intramuscular injection were safe and well-tolerated. Respiratory adverse events were infrequent, mild, transient and similar among groups (Table 3). Intramuscular injection was associated with a mild local injection site reaction in two participants. Systemic adverse events were also infrequent, mild, transient and similar among groups (Table 3). One participant who received low dose aerosol vaccine developed genital lesions consistent with primary HSV-1 infection the day following the week 2 bronchoscopy, which resolved without complication with oral valacyclovir, and one participant developed plantar fasciitis on day 13 following vaccination which was attributed to mechanical strain and resolved with acetominophen. There were no grade 3 or 4 adverse events reported nor any serious adverse events.

**Table 3.** Adverse events

| Adverse event | Low dose (n=11) | % | High dose (n=12) | % | IM (n=9) | % |
| --- | --- | --- | --- | --- | --- | --- |
| Injection site reaction |  |  |  |  |  |  |
| Local pain | n/a | n/a | n/a | n/a | 0 | 0 |
| Redness | n/a | n/a | n/a | n/a | 1 | 11.1 |
| Swelling | n/a | n/a | n/a | n/a | 1 | 11.1 |
| Cough | 2 | 18.1 | 2 | 16.6 | 4 | 44.4 |
| Runny nose | 3 | 27.7 | 2 | 16.6 | 2 | 22.2 |
| Fatigue | 4 | 36.3 | 3 | 25.0 | 3 | 33.3 |
| Myalgia | 3 | 27.7 | 0 | 0 | 0 | 0 |
| Headache | 3 | 27.7 | 7 | 58.3 | 2 | 22.2 |
| Joint pain | 1 | 9.0 | 0 | 0 | 1 | 11.1 |
| Chest pain | 1 | 9.0 | 0 | 0 | 0 | 22.2 |
| Sneezing | 1 | 9.0 | 1 | 8.3 | 2 | 0 |
| Sinus pain | 1 | 9.0 | 0 | 0 | 0 | 0 |
| Abdominal pain | 1 | 9.0 | 1 | 8.3 | 1 | 11.1 |
| Shortness of breath | 0 | 0 | 1 | 8.3 | 0 | 0 |
| Nose bleed | 1 | 9.0 | 0 | 0 | 0 | 0 |
| Itchy throat | 0 | 0 | 1 | 8.3 | 0 | 0 |
| Shivers | 2 | 18.1 | 0 | 0 | 0 | 0 |
| Dizziness | 0 | 0 | 0 | 0 | 2 | 22.2 |

There were no clinically significant abnormalities of laboratory tests at weeks 2, 4 and 12 following vaccination. Follow-up respiratory functional determinations FEV1 and FVC were similar to baseline values in all participants across all three groups (Fig S2-A/B).

Bronchoscopy and bronchoalveolar lavage were generally well tolerated in all participants. As expected, in some participants the procedures were associated with mild cough, sore throat, low-grade fever, headache and a transient drop in FEV1. The appearance of the bronchial mucosa was judged as normal in all participants at each time point. Adequate bronchoalveolar lavage fluid (BALF) volumes were obtained following bronchoalveolar lavage and on average 10-20 million total cells were obtained.

### Aerosol AdHu5Ag85A vaccination induces robust and sustainable Th1 responses in the airway

Bronchoalveolar lavage was obtained successfully from all participants with a median return volume of 87.5 ml (IQR: 72.5-98) from a total of 160 ml saline instilled and a median total cell number of 0.14 million/ml BALF (IQR: 0.1-0.2). Cellularity in the airway significantly increased 2wk after both low dose (LD) and high dose (HD) aerosol vaccination and in LD aerosol group, cellularity remained significantly heightened up to 8wk post-vaccination compared with baseline (Fig S2-C). Both LD and HD aerosol vaccination led to a transient reduction in airway macrophages, but the lymphocyte counts significantly increased only in LD cohort (Fig S2-D/E). Importantly, both neutrophils and epithelial cells in the airway remained either absent or unaltered following aerosol vaccination (Fig S2-D/E), indicating no significant airway inflammation. In comparison, there were no marked changes in total cellularity and any leukocyte subsets in the airway after intramuscular (IM) vaccination (Fig S2-C/F).

Evaluation of Th1 responses in the airways (BALF) cells by intracellular cytokine immunostaining showed that both LD and HD aerosol AdHu5Ag85A markedly increased Ag85A peptide pool (Ag85A p. pool)-specific or reactive, IFNγ-, TNFα- and/or IL-2-producing CD4 T cells in the airways at 2wk after vaccination compared with the respective baseline responses (Fig 2A/B/C). On average, the total Ag-specific cytokine-producing CD4 T cells represented ∼25% of all CD4 T cells at 2wk post-LD or HD aerosol and they remained significantly elevated up to 8wk in the airways of LD cohort (Fig 2A/B). In contrast, IM AdHu5Ag85A vaccination failed to induce Ag-specific CD4 T cells in the airways (Fig 2A). Compared with CD4 T cells, although the levels of airway CD8 T cell responses were much smaller, they were significantly increased at both 2 and 8wk, particularly following LD aerosol vaccination (Fig 2D). Of interest, there was also a small but increased number of CD8 T cells at 2wk after IM vaccination.

**Figure 2.**
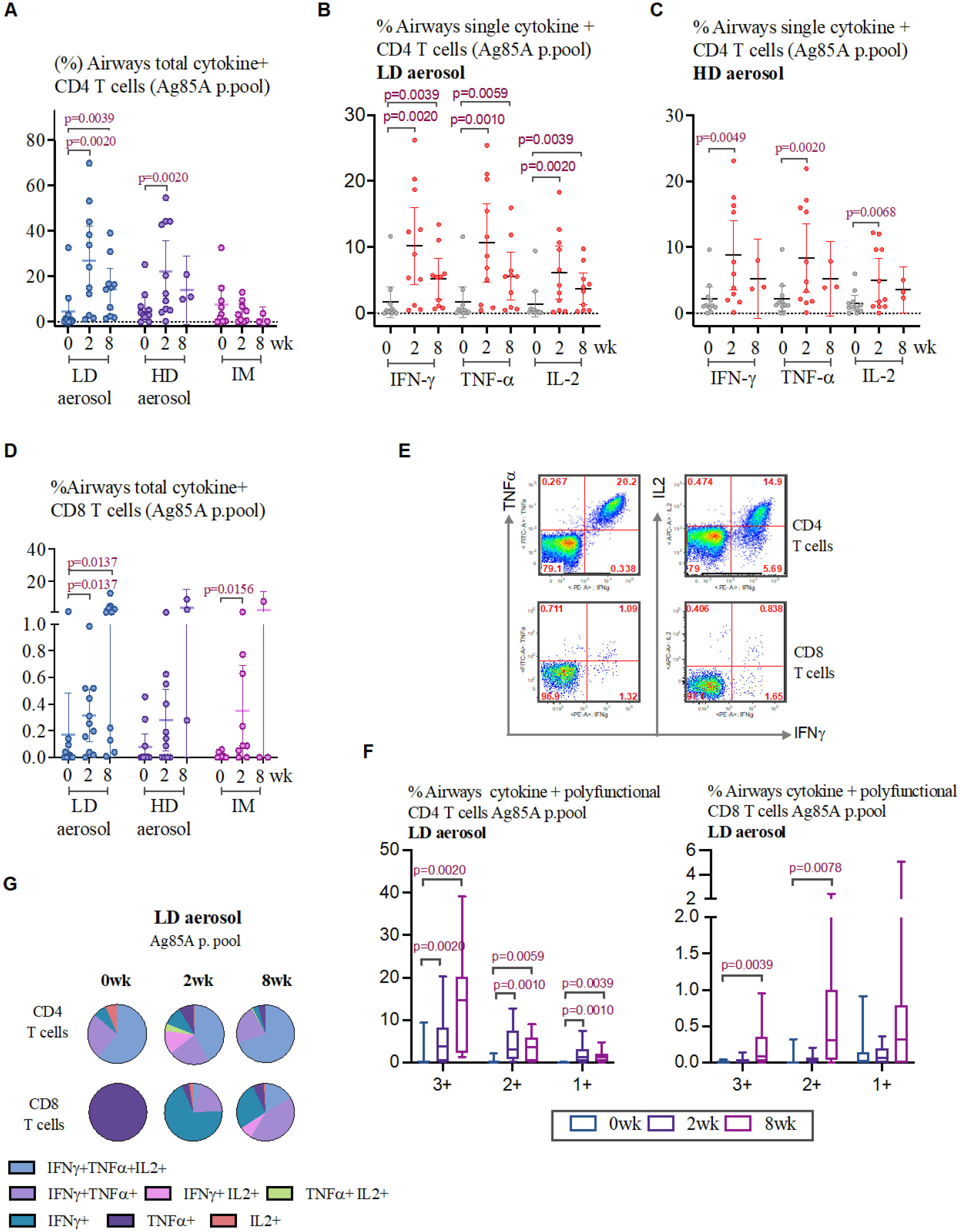
Induction of multifunctional T cells in the airways following aerosol or intramuscular vaccination. **A**) Frequencies of airways antigen-specific combined total cytokine-producing CD4 T cells at various timepoints in LD aerosol, HD aerosol and IM cohorts. **B**) Frequencies of airways single cytokine-producing CD4 T cells at various timepoints in LD aerosol cohort. **C**) Frequencies of airways single cytokine-producing CD4 T cells at various timepoints in HD aerosol cohort. D) Frequencies of airways antigen-specific combined total cytokine-producing CD8 T cells at various timepoints in LD aerosol, HD aerosol and IM cohorts. **E**) Representative dotplots of airways CD4 and CD8 T cells expressing IFNγ, TNFα and IL-2 at wk2 from LD aerosol participants. **F**) Frequencies of airways polyfunctional (triple/3+, double/2+ and single/1 cytokine-positive) antigen-specific CD4 and CD8 T cells at various timepoints in LD aerosol group. **G**) Median proportions displayed in pie chart of antigen-specific airways CD4 and CD8 T cells expressing a specific single or combination of two or three cytokines at various timepoints in LD aerosol group.

Analysis of polyfunctionality of vaccine-activated CD4 and CD8 T cells reactive to Ag85A in the airways revealed that LD aerosol vaccination led to induction of higher magnitude of CD4 T cells that co-expressed IFNγ, TNFα and IL-2 (3+) and any of two cytokines (2+) than those producing single cytokine (1+) (Fig 2E/F). Importantly, polyfunctional CD4 T cells remained significantly increased over the baseline up to 8wks. Similarly, LD aerosol vaccination also significantly increased the polyfunctional CD8 T cells, particularly at 8wks, in the airways, though at much lower overall magnitude compared to CD4 T cells (Fig 2F). In comparison, HD aerosol vaccination led to significantly increased polyfunctional CD4 but not CD8 T cells reactive to Ag85A (Fig S3-A/B).

Given that the LD aerosol AdHu5Ag85A was consistently highly immunogenic, we profiled the polyfunctional CD4 and CD8 T cells in the airways of LD cohort in greater detail. While at 2wk, a greater proportion of Ag85A-reactive CD4 T cells were polyfunctional (IFNγ+TNFα+IL-2+, IFNγ+TNFα+ or IFNγ+IL-2+) with some of them also being single cytokine producers, at 8wk the vast majority of them (>95%) became polyfunctional (Fig 2G). In comparison, most of the CD8 T cells at 2wk were single cytokine producers (TNFα+) but at 8wk the majority of them turned to be polyfunctional, mostly being IFNγ+TNFα+ (Fig 2G).

Since the trial participants were previously BCG-vaccinated, we examined the overall T cell reactivity to stimulation with multi-mycobacterial antigens. We found considerable CD4 T cells present in the airways to be reactive to a cocktail of mycobacterial antigens even prior to vaccination and they remained unaltered after LD, HD or IM vaccination (Fig S3-C). These BCG-specific CD4 T cells in the airways of LD group were mostly polyfunctional (Fig S3-D/E). Similarly, small numbers of pre-existing BCG-specific CD8 T cells in the airways were not altered by aerosol or IM vaccination (Fig S3-F).

The above data suggest that inhaled aerosol, but not intramuscular, AdHu5Ag85A vaccination can induce robust antigen-specific T cell responses within the respiratory tract. Furthermore, a low-dose (1×10^6^ PFU) aerosol vaccination is superior to a high dose (2×10^6^ PFU) aerosol in inducing robust and sustainable respiratory mucosal immunity. The mucosal responses induced by AdHu5Ag85A vaccine are predominantly polyfunctional CD4 T cells in nature with some levels of polyfunctional CD8 T cells. The pre-existing CD4 T cells of multi-mycobacterial antigen specificities in the airway of BCG-vaccinated trial participants were not significantly impacted by AdHu5Ag85A aerosol vaccination.

### Aerosol vaccination induces airway-resident memory T cells (T_RM_) expressing the lung homing molecule α4β1 integrin

Lung tissue-resident memory T cells are critical to protective mucosal immunity ^19^. Hence, we next determined whether antigen-specific T cells induced by aerosol AdHu5Ag85A vaccination were of tissue-resident memory phenotype and compared them to those by IM vaccination. BALF cells obtained before and at select timepoints post-vaccination were stimulated with Ag85A p. pool and immunostained for co-expression of two key T_RM_ surface markers CD69 and CD103 by antigen-specific IFNγ-producing CD4 or CD8 T cells (Fig 3A). Marked increases in Ag85A-specific IFN-γ+ CD4 and CD8 T cells co-expressing CD69 and CD103 were seen only in the airway of LD and HD aerosol vaccine groups, but not in IM group (Fig 3B/C). Although T_RM_ increases at 8wk post-aerosol vaccination compared to the baseline were only marginally statistically significant (95% CI) probably due to small sample sizes, remarkable proportions of Ag85A-specific CD4 T cells (∼20%) and CD8 T cells (∼54%) present in the airways of aerosol vaccine groups were T_RM_ (Fig 3D). As expected, there was no detectable antigen-specific T_RM_ in the peripheral blood before and after vaccination.

**Figure 3.**
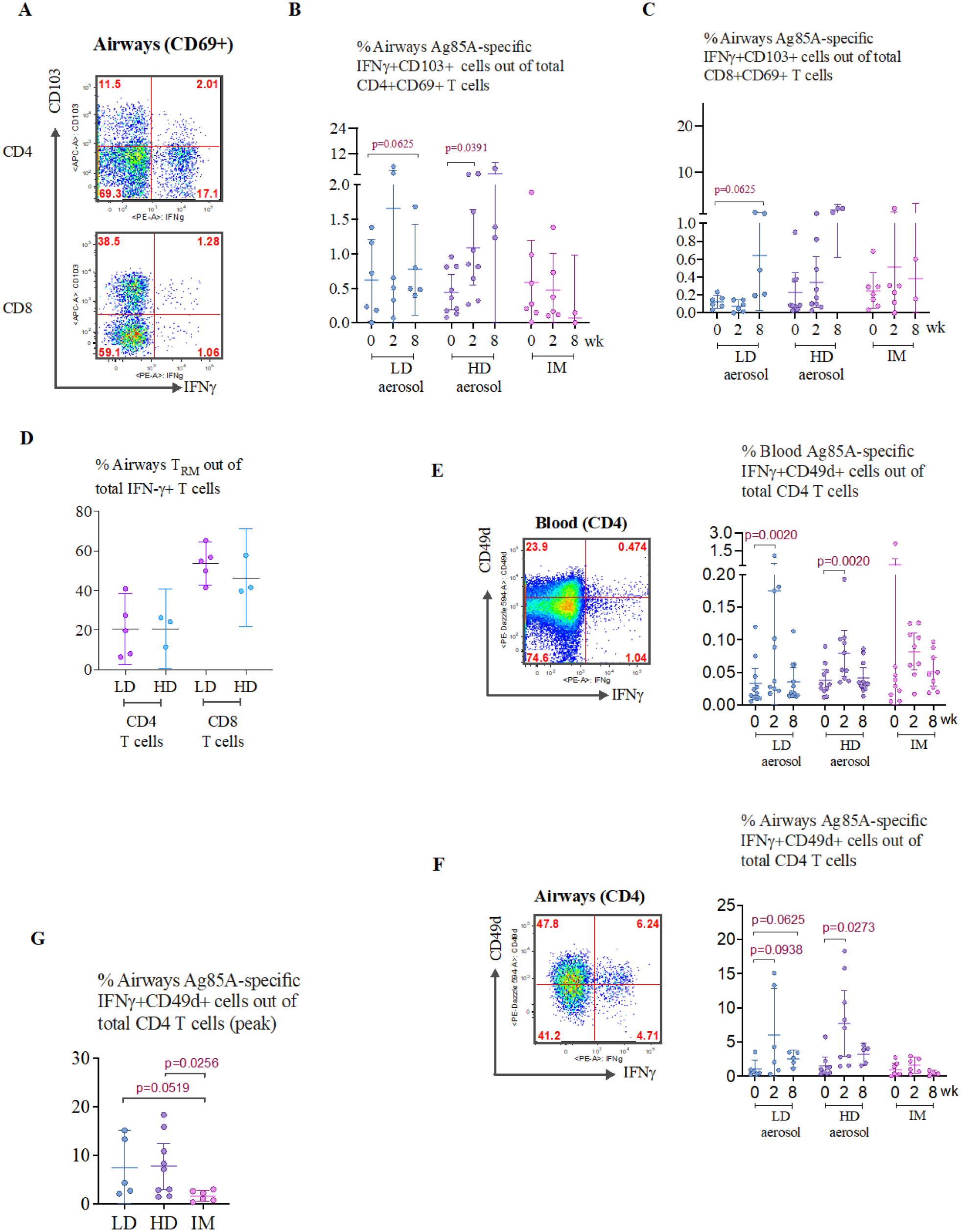
Induction of airway tissue-resident memory T cells (T_RM_) following aerosol or intramuscular vaccination. **A**) Representative dotplots of airways antigen-specific CD4 and CD8 T_RM_ at wk2 in LD aerosol participants. **B**) Frequencies of airways antigen-specific IFNγ+CD4 T_RM_ co-expressing CD69 and CD103 surface markers at various timepoints in LD aerosol, HD aerosol and IM vaccine cohorts. **C**) Frequencies of airways antigen-specific IFNγ+CD8 T_RM_ co-expressing CD69 and CD103 at various timepoints in LD aerosol, HD aerosol and IM vaccine cohorts. **D**) Comparison of frequencies of airways antigen-specific CD4 and CD8 T_RM_ co-expressing CD69 and CD103 at 8wk post-LD and HD aerosol vaccination. **E**) Representative dotplots of peripheral blood antigen-specific IFNγ+CD4 T cells expressing CD49d at wk2 from LD aerosol participants, and frequencies of circulating antigen-specific CD4 T cells expressing CD49d at various timepoints in LD aerosol, HD aerosol and IM vaccine cohorts. **F**) Representative dotplots of airways antigen-specific IFNγ+CD4 T cells expressing CD49d at wk2 from LD aerosol participants, and frequencies of airways antigen-specific CD4 T cells expressing CD49d at various timepoints in LD aerosol, HD aerosol and IM vaccine cohorts. **G**) Comparison of frequencies of airways antigen-specific IFNγ+CD4 T cells co-expressing CD49d at the peak timepoint in LD aerosol, HD aerosol and IM vaccine cohorts.

We also studied T cell surface expression of α4β1 integrin (VLA-4; or CD49d for α4), known to be expressed on memory CD4 T cells in human airways ^20^. Since CD49d may be involved in the homing of circulating T cells to the airway, we first examined CD49d expression on Ag85A-specific CD4 T cells in the circulation. There were small but significantly increased frequencies of circulating CD49d-expressing IFNγ+CD4 T cells, particularly at 2wk following LD or HD aerosol vaccination (Fig 3E). In comparison, there were much greater frequencies of CD49d-expressing IFNγ+CD4 T cells (out of total CD4 T cells) in the airways induced by aerosol vaccination (Fig 3F), compared to their frequencies in the circulation (Fig 3E) and in contrast with the lack of such T cells in the airways of IM group (Fig 3G). In fact, the majority of Ag85A-specifc CD4 T cells in the airways of LD and HD groups expressed CD49d (57% and 74%, respectively). The data together indicate that aerosol AdHu5Ag85A vaccination, but not IM route of vaccination, is uniquely capable of inducing antigen-specific T cells in the airways endowed with respiratory mucosal homing and T_RM_ properties.

Since besides mucosal adaptive immunity, respiratory delivery of AdHu5Ag85A vaccine in experimental animals induced a trained phenotype in airway macrophages ^6, 21^, we examined whether aerosol vaccination could also alter the immune property of human alveolar macrophages (AM). To this end, we elected to examine the transcriptomics of BALF cells obtained from 5 participants before (wk0) and after (wk8) LD aerosol vaccination. Before RNA isolation, the cells, upon revival from frozen stock, were enriched for AM and cultured with or without stimulation with *M.tb* lysates and transcriptionally profiled by RNAseq analysis. Principal component analysis (PCA) revealed that unstimulated (US) and stimulated (S) AM populations were separated away from each other (Fig S4-A). We then identified the differentially expressed genes (DEGs) by comparing wk0- and wk8-stimulated AM with respective unstimulated AM. A total of 2,726 genes were differentially expressed upon stimulation in pairwise analysis of which 1667 genes (61%) were shared between the baseline (wk0) Group 3/1 and aerosol vaccine (wk8) Group 4/2 (Fig S4-B). As expected, the shared genes were significantly enriched in biological processes associated with immune response and regulation of cell death (Fig S4-C). Further by pairwise analysis, we identified 194 and 426 genes uniquely upregulated and downregulated, respectively, in stimulated aerosol (Group 4) AM (Fig S4-D). The uniquely up-regulated gen<u>e</u>s in stimulated wk8 aerosol AM showed enrichment in a number of biological processes including response to anoxia (OXTR, CTGF), inflammatory response to antigenic stimuli (IL2RA, IL1B, IL20RB), tyrosine phosphorylation of STAT protein (IFNG, F2R, OSM), regulation of IL-10 production (CD83, IRF4, IL20RB, IDO1), response to IL-1 (RIPK2, SRC, IRAK2, IL1R1, XYLT1, RELA) <u>and</u> histone demethylation (KDM6B, KDM5B, KDM1A, KDM7A, JMJD6) (Fig S4-E). In comparison, the uniquely down-regulated genes in wk8 aerosol AM did not appear significantly enriched for any biological processes. These data suggest that LD aerosol vaccination leads to persisting transcriptional changes in airway-resident alveolar macrophages poised for defense responses.

### Aerosol vaccination induces levels of systemic Th1 responses comparable to intramuscular vaccination

Assessment of overall antigen-specific reactivity of T cells in the circulation before and after vaccination by using whole blood samples incubated with Ag85A peptides indicated that both aerosol, particularly LD aerosol, and IM AdHu5Ag85A vaccination induced significant systemic immune responses as shown by raised IFNγ, TNFα and IL-2 levels in plasma (Fig 4A/B/C). Area under curve (AUC) analysis, which reflects the overall magnitude of responses, did not differ between LD aerosol and IM groups in cytokine production in response to Ag85A p. pool stimulation (IFNγ p=0.0910; TNFα p=0.6207; IL-2 p=0.8703). However, IM vaccine-induced systemic T cell responses appeared to remain significantly increased over a longer duration (Fig 4C). In comparison, HD aerosol group had significantly lower IFNγ production than IM group (AUC compared to IM-p= 0.0117) whereas they did not differ from each other in the production of TNFα and IL-2 (Fig 4B/C).

**Figure 4.**
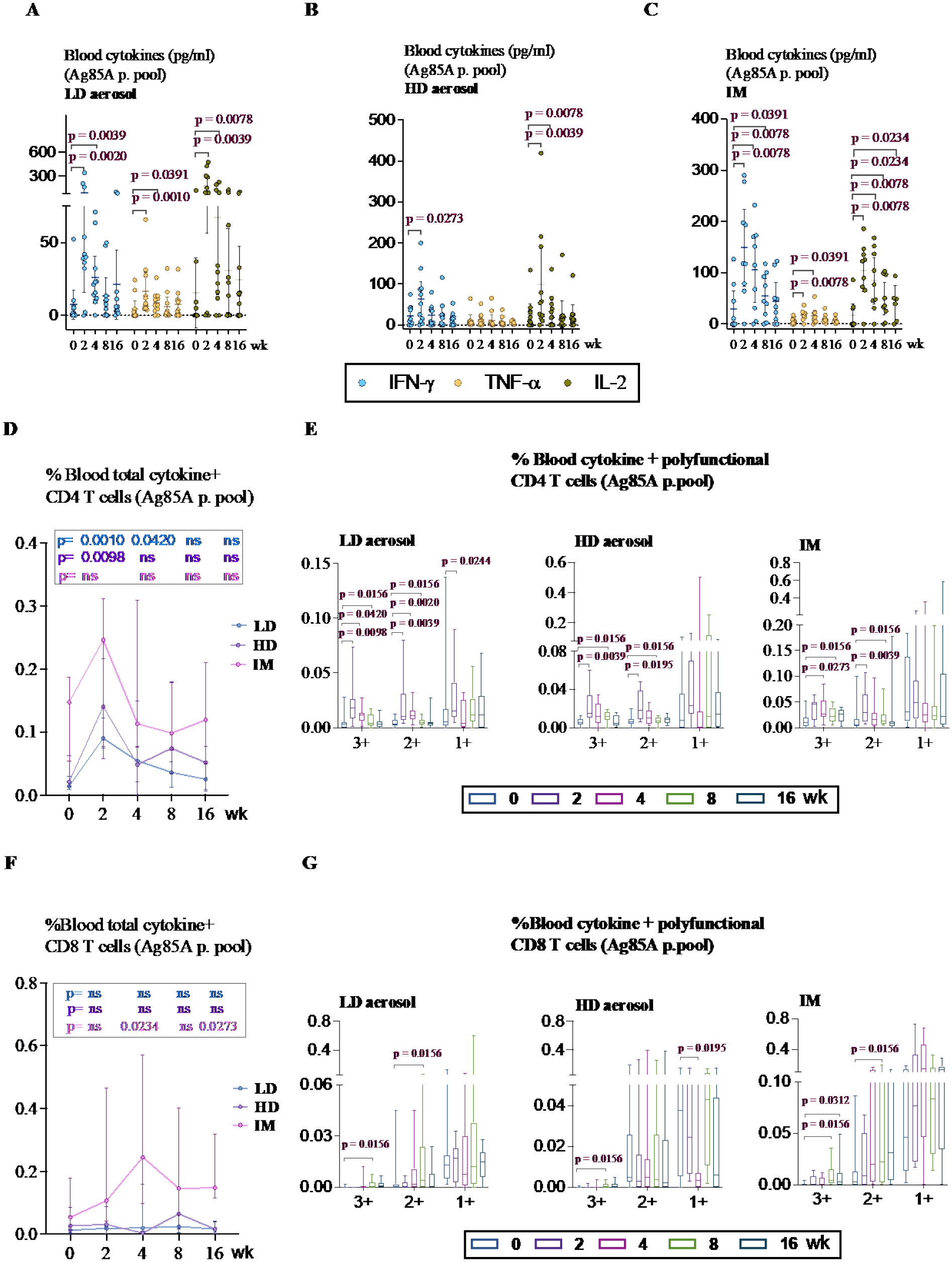
Induction of antigen-specific T cell responses in the peripheral blood following aerosol or intramuscular vaccination. **A**),**B**),**C**) Antigen-specific cytokine production in whole blood culture at various timepoints post-LD aerosol, HD aerosol and IM vaccine groups. The measurements were subtracted from unstimulated control values. **D**) Frequencies of peripheral blood antigen-specific combined total cytokine-producing CD4 T cells at various timepoints in LD aerosol, HD aerosol and IM cohorts. **E**) Frequencies of peripheral blood polyfunctional (triple/3+, double/2+ and single/1 cytokine-positive) antigen-specific CD4 T cells at various timepoints in LD aerosol, HD aerosol and IM vaccine groups. **F**) Frequencies of peripheral blood antigen-specific combined total cytokine-producing CD8 T cells at various timepoints in LD aerosol, HD aerosol and IM groups. **G**) Frequencies of peripheral blood polyfunctional (triple/3+, double/2+ and single/1 cytokine-positive) antigen-specific CD8 T cells at various timepoints in LD aerosol, HD aerosol and IM groups.

Further examination of relative activation of CD4 and CD8 T cells by aerosol and IM vaccinations using intracellular cytokine staining revealed that compared to the respective baseline, aerosol vaccination activated the circulating Ag85A-specific CD4 T cells to significant levels while IM vaccination moderately increased such responses (Fig 4D). However, the overall magnitude of responses did not differ significantly between aerosol and IM groups (AUC: LD aerosol-p=0.1961; HD aerosol-p=0.3545 compared to IM). Both LD/HD aerosol and IM vaccination also significantly increased Ag85A-specific polyfunctional CD4 T cells co-expressing three (3+) or any two (2+) cytokines in the circulation (Fig 4E). Consistent with the airway Ag85A-specific CD4 T cell responses (Fig 2A/B/C) in both LD and HD aerosol groups, circulating polyfunctional CD4 T cells also generally peaked at 2wk post-vaccination and remained significantly increased up to 8wk (Fig 4E). In comparison, 3+ polyfunctional CD4 T cells in IM group significantly increased at 4wk and remained increased up to 8wk (Fig 4E). The overall magnitude of circulating 3+ polyfunctional CD4 T cells in IM group was however significantly higher than those in aerosol groups (AUC: LD aerosol, p=0.0144 and HD aerosol, p=0.0393 compared to IM). Circulating 2+ polyfunctional CD4 T cells did not differ significantly between these groups (AUC not significantly different). Consistent with our previous observation ^8^, besides its activating effects on circulating CD4 T cells, IM vaccination also significantly increased Ag85A-specific CD8 T cells up to 16wk (Fig 4F). By comparison, aerosol vaccination minimally induced such CD8 T cells in the circulation (Fig 4F). Compared to circulating CD4 T cells, similar to the overall kinetics of total cytokine+ CD8 T cells (Fig 4F), circulating Ag85A-specific polyfunctional CD8 T cells peaked behind the peak CD4 T cell responses in all vaccine groups (Fig 4G).

The kinetics of polyfunctional profiles of circulating CD4 T cells were further examined in greater detail with a focus on LD aerosol vaccine group and its comparison with IM group. There existed considerable differences in the polyfunctional profile of circulating Ag85A-specific CD4 T cells between LD aerosol and IM groups (Fig S5-A). In LD aerosol group, the proportion of IFNγ+TNFα+IL-2+ progressively shrank and at 16wk, ∼ 75% of the population were TNFα+IL-2+ and IFNγ+TNF-α+ together with single TNFα+ CD4 T cells. In comparison, in IM group the proportion of IFNγ+TNFα+IL-2+ progressively expanded, constituting ∼ 75% of the population at 16wk (Fig S5-A).

Upon examination of circulating BCG-specific CD4 T cells (reactive to *M.tb*CF+rAg85A stimulation), we found that they were not strikingly increased in aerosol and IM vaccine groups although the trend was higher in IM group (Fig S5-B/C) and AUC values did not differ significantly between aerosol and intramuscular groups (LD aerosol, p=0.0870; HD aerosol, p=0.2666 compared to IM). However, LD and HD aerosol vaccination had a significant enhancing effect on the polyfunctionality of pre-existing circulating BCG-specific CD4 T cells (Fig S5-C). Similar to BCG-specific circulating CD4 T cells (Fig S5-B), BCG-specific circulating CD8 T cells were not significantly increased by either aerosol or IM vaccination (Fig S5-D).

These data together indicate that besides markedly induced mucosal T cell immunity (Fig 2/3), respiratory mucosal vaccination via inhaled aerosol, particularly LD aerosol, can also induce systemic polyfunctional CD4 T cell responses at levels comparable to those by IM route of vaccination in previously BCG-vaccinated humans.

### Pre-existing anti-AdHu5 antibodies have minimal effects on the immunogenicity of aerosol AdHu5Ag85A vaccination

The high prevalence of circulating pre-existing antibodies (Ab) against AdHu5 in human populations may negatively impact on the potency of AdHu5-vectored vaccines following intramuscular administration ^22^. However, little is known about its effect on the potency of respiratory mucosal-delivered AdHu5-vectored vaccine. To address this question, we first examined the levels of AdHu5-specific total immunoglobulin G (IgG) in the circulation and airways (BALF) before and after vaccination (wk 0 vs 4 in circulation; wk 0 vs. 8 in BALF). In keeping with our previous findings ^8^, there were significant levels of pre-existing circulating AdHu5-specific total IgG in most of the trial participants (10^4^-10^5^), and the levels were comparable between the groups (using Kruskal-Wallis test p=0.2048) (Table 4). These titres significantly increased after HD aerosol or IM AdHu5Ag85A vaccination but not after LD aerosol vaccination. In comparison, pre-existing levels of anti-AdHu5 total IgG in the airways were 1-1.5 log less than the levels in the circulation and were comparable between groups (using Kruskal-Wallis test p=0.2048). Of interest, LD and HD aerosol as well as IM vaccination did not alter the pre-existing anti-AdHu5 total IgG levels in the airways (Table 4) but the data from HD aerosol and IM groups should be interpreted with caution due to the small sample size at 8wk.

**Table 4.** Anti-Ad5 antibody titers before and after LD aerosol, HD aerosol, and IM vaccination

| Sample | Ad5 humoral immunity | Study time (wk) |  | LD aerosol |  | HD aerosol |  | IM |
| --- | --- | --- | --- | --- | --- | --- | --- | --- |
|  |  |  | N |  | N |  | N |  |
| Serum | Anti-Ad5 Total IgG titre | 0 | 11 | 9513<br>(700, 115520) | 11 | 5695<br>(932, 39080) | 9 | 14257<br>(4974, 67200) |
|  |  | 4 | 11 | 4974<br>(611, 326600) | 11 | 9384*<br>(1161, 68960)<br>**P=0.0010** | 9 | 20350*<br>(5744, 61200)<br>**P=0.0017** |
|  | Ad5 neutralizing antibody titre | 0 | 11 | 22<br>(2, 7937) | 11 | 130<br>(2, 7519) | 9 | 203<br>(2, 11110) |
|  |  | 4 | 11 | 32<br>(3, 7692) | 11 | 134<br>(2, 7634) | 9 | 2311*<br>(25, 19231)<br>**P=0.0039** |
| BALF | Anti-Ad5 Total IgG titre | 0 | 11 | 469<br>(222, 2855) | 11 | 522<br>(84, 3441) | 9 | 281<br>(24, 3290) |
|  |  | 8 | 10 | 502<br>(115, 1188) | 3 | 213<br>(98, 859) | 3 | 147<br>(84, 194) |
|  | Ad5 neutralizing antibody titre | 0 | 11 | 2.13<br>(1, 55) | 11 | 13<br>(1, 93) | 9 | 5.9<br>(1-139) |
|  |  | 8 | 10 | 2.8<br>(1, 46) | 3 | 14<br>(1-79) | 3 | 6.4<br>(1-30) |

Because the total anti-AdHu5 Ab titres may not always correlate with AdHu5-neutralizing capacity in the circulation ^8^, we further assessed the AdHu5-neutralizing Ab (nAb) titres before and after vaccination in the circulation and airways by using a bioassay. The pre-existing AdHu5-nAb titres in the circulation were comparable between groups (using Kruskal-Wallis test p=0.3588) with 27%, 54% and 66% of participants in LD, HD and IM groups having >10^2^ AdHu5-nAb titres, respectively. Of interest, while IM vaccination with AdHu5Ag85A significantly increased the circulating AdHu5-nAb titers by an average of 1.5 logs, LD or HD aerosol vaccination had no such effect (Table 4). On the other hand, similar to total anti-AdHu5 IgG levels, pre-existing AdHu5-nAb titers in the airways were ∼1 log less than those in the circulation (Table 4). Of importance, 63%, 36% and 33% of participants in LD, HD and IM groups, respectively, had no detectable baseline AdHu5-nAb titers in their airways which remained unaltered following vaccination (Table 4). We further found a significant positive correlation between AdHu5-nAb and total AdHu5 IgG titres both in the circulation and airways (Fig S6-A/B).

Given that many of the trial participants had moderate to significant levels of AdHu5-nAb titers in the circulation and ∼50% of them also had a small but detectable level of pre-existing AdHu5-nAb titres in the airways, we next examined whether such nAbs present in the airways and blood may have negatively impacted the immunopotency of LD aerosol and IM vaccination, respectively. To this end, % airways or blood total cytokine+ Ag85A-specific CD4 T cells at the peak response time (2 weeks post-vaccination) for individual participants was plotted against corresponding pre-existing AdHu5-nAb titre and Spearman rank correlation test was performed. There was no significant correlation between pre-existing airways AdHu5-nAb titers and the magnitude of vaccine-induced CD4 (Fig S6-C) and CD8 (Fig S6-D) T cell responses in the airways following LD aerosol vaccination. Of note, one participant who hardly responded to aerosol vaccine did have the highest neutralization titers in the cohort (Fig S6-C). On the other hand, consistent with our previous observation ^8^, there was no significant correlation between pre-existing circulating AdHu5-nAb titers and the magnitude of antigen-specific CD4 (Fig S6-E) and CD8 (Fig S6-F) T cell responses in the blood following IM vaccination. The above data together suggest that while there is high prevalence of pre-existing circulating anti-AdHu5 nAb in humans enrolled in our study, most of trial participants have either undetectable or very low levels of pre-existing anti-AdHu5 nAb titers in the airways. IM AdHu5Ag85A vaccination increases AdHu5 nAb titers in the circulation whereas aerosol vaccination does not do so either in the airways or in the circulation. Furthermore, overall, the presence of AdHu5 nAb does not seem to have a significant impact on vaccine immunogenicity.

## Discussion

This represents the first clinical study to have fully characterized the method to deliver aerosolized adenoviral-vectored vaccine to human lungs and to demonstrate its superiority in inducing respiratory mucosal immunity over the intramuscular injection.

Both low and high aerosol doses of AdHu5Ag85A were safe and well-tolerated. Respiratory adverse events were rare, mild, transient and similar among groups and the respiratory function determined by FEV1 and FVC were within normal limits. Systemic adverse events were also similarly rare, mild and transient. There were no grade 3 or 4 adverse events reported nor any serious adverse events. There were no clinically significant abnormalities of routine laboratory tests at weeks 2, 4 and 12 following either aerosol or IM vaccination.

Our finding that respiratory mucosal vaccination via inhaled aerosol delivery, but not the parenteral intramuscular vaccination, is capable of induction of robust respiratory mucosal immunity in human lungs is consistent with the well-established knowledge from animal models of TB and other respiratory infections ^3, 23–25^. While both the low and high aerosol doses were found to be safe, our findings support the low-dose aerosol, rather than the high-dose, to be optimal as it led to more consistently increased T cell responses in the airway. Of note, this low aerosol dose (10^6^ PFU) was 100 times smaller than the IM dose (10^8^ PFU) in our study. The remarkable immunogenicity of a rather small dose of a human serotype 5 Ad (AdHu5)-vectored vaccine at human respiratory mucosa was observed despite the common knowledge that the majority of humans have varying degrees of pre-existing antibodies against AdHu5 ^22, 26^. The superiority of AdHu5-vectored vaccine delivered via aerosol, as opposed to IM injection, in inducing respiratory mucosal immunity, suggests the respiratory tract to be an immune privileged site in terms of anti-AdHu5 nAb. Indeed, we find that ∼50% of our trial participants have no detectable pre-existing nAb titers and the rest of them have rather low pre-existing nAb titers in the airways, contrasting to readily measurable pre-existing nAb titers in the blood of most of the trial participants. Such compartmentalized prevalence of anti-AdHu5 Ab in the circulation, but not in the airways of humans, is also reported in a previous study ^27^. Consideration of the relative prevalence of anti-human Ad nAb is important to the design of human Ad-based vaccine strategies ^15, 22, 28^. In this regard, although probably due to relatively small sample size, we did not find a significant negative correlation between AdHu5 nAb titers and T cell responses in the peripheral blood following IM injection of AdHu5Ag85A vaccine, a strong negative correlation has been seen in larger clinical trials with AdHu5-based vaccines ^14, 29^. Also, for the first time, we find that aerosol delivery of an AdHu5-vectored vaccine did not enhance AdHu5 nAb in the airways whereas the IM route of vaccination significantly enhanced nAb titers in the blood. This represents an additional advantage of inhaled aerosol delivery of Ad-vectored vaccine, suggesting the feasibility of repeated aerosol deliveries of such vaccines for undiminished immunopotency. Thus, our findings together suggest that besides its unique strength and advantage in inducing much desired respiratory mucosal immunity and repeatability, the inhaled aerosol Ad-based vaccine strategy is pain/needle-free and requires only a small dose to be effective, helping with global vaccine distribution when vaccine supplies may be limited.

Pulmonary TB has remained a major global health issue. Despite great strides made in the past couple of decades to develop improved TB vaccine strategies, BCG, a century-old TB vaccine, still continues to be used worldwide ^30^. To effectively control the global TB epidemic, new, unconventional vaccine approaches such as respiratory mucosal vaccine strategies are likely required ^2, 3, 5, 24^. Respiratory mucosal immunization strategies with appropriately chosen vaccine platforms are adept at inducing the all-round protective immunity consisting of humoral immunity, tissue-resident memory T cells, and trained innate immunity ^3^. The scientists at Jenner Institute of Oxford University pioneered an inhaled aerosol method to deliver an MVA-vectored TB vaccine to human respiratory tract and found much improved T cell responses in the airways over that by intradermal injection ^10, 12^. Of note, a different aerosol device was used, and the aerosol particles and delivery efficiency were not characterized in these studies. Furthermore, since in these studies, bronchoscopy was performed only once to assess the local mucosal immune responses at 1week post-vaccination, it is unclear whether aerosol vaccine-activated effector T cells may persist, becoming long-term memory T cells in the airways. In comparison, in our current study, not only were the aerosol droplets and delivery efficiency fully characterized, but bronchoscopy was performed before vaccination and twice at 2 and 8 weeks post-vaccination. Since, as shown in our current study and by others ^31^, the airways of previously BCG-vaccinated or PPD-positive humans harbor mycobacterial antigen-specific T cells at baseline, we believe that bronchoscopy carried out before and after vaccination is needed to help differentiate the authenticity of vaccine-induced T cell responses. Based on immune analysis of the cells harvested via 8-week post-aerosol bronchoscopy, our study for the first time offers the critical evidence that inhaled aerosol viral-vectored vaccine can induce persisting tissue-resident memory T cells within human respiratory tract. Using RNAseq, our study also reveals the lasting transcriptional changes in airway macrophages, indicative of trained innate immunity, following aerosol vaccination. We have recently reported that in preclinical models, respiratory mucosal-delivered AdHu5-vectored TB vaccine induces lasting memory airway macrophages capable of trained innate immunity against *M.tb* and unrelated bacterial pathogens ^6, 21^. Incorporating the ability of a vaccine vector to induce tissue-resident trained innate immunity alongside tissue-resident adaptive immunity into the design of TB vaccine strategies may provide the best possible protection ^3, 5^.

The well-characterized inhaled aerosol vaccine technology and its superiority to induce respiratory mucosal immunity demonstrated in our current study also offers a foundation and the important proof of concept for developing the next-generation COVID-19 vaccine strategies. Indeed, the effective global control of COVID-19 via the roll-out of the first-generation vaccines has met with threats from emerging variants of concern, dwindling vaccine-induced immunity, and increasing break-through infections ^16^. Of note, several approved first-generation COVID-19 vaccines are adenoviral-vectored but they are all administered via intramuscular injection ^15^. Although a recent study has shown that it is safe and well-tolerated to deliver via inhaled aerosol an AdHu5-vectored COVID-19 vaccine into human respiratory tract ^14^, there are a number of critical drawbacks associated with this study. This study did not evaluate the local respiratory mucosal immune responses following aerosol and intramuscular vaccination and it is unclear that how much aerosolized biologically active vaccine was deposited into human respiratory tract. Furthermore, in this study, 2-repeated high aerosol doses (up to 2×10^10^ vp/dose) were compared with an IM prime-aerosol boost regimen, leaving the question open whether 2-repeated aerosol doses are scientifically justified or is it counterproductive. As we have shown in our current study, the respiratory tract is an immunologically highly conducive site for adenoviral-vectored immunization and a rather small single aerosol dose (∼3×10^7^ vp) is able to induce high frequencies of antigen-specific tissue-resident memory T cells in the airways. We further show that a relatively small increase in aerosol dose did not translate to greater T cell responses, but rather, it caused inconsistent immune responses probably due to immune overactivation.

In conclusion, our current study has provided technological details for safely and efficiently delivering aerosolized adenoviral-vector vaccine into human respiratory tract and demonstrated its superiority to induce respiratory mucosal immunity over parenteral intramuscular injection. Our findings will foster the development of effective vaccine strategies against not only TB but also other respiratory infections including COVID-19.

## Data Availability

All data related to this clinical vaccine trial will be made available

https://data.mendeley.com/

## Contributors

FS, ZX, GMG, MJ designed the trial. MBD, MCM, MRT, ZX, FS characterized the aerosol device and aerosol droplets. BDL, MCM took charge of vaccine GMP manufacturing. MJ, DKF, SA, AZ, AD performed immunological assays and data analysis. EA, KJH, RFS, RC, PMO, MK, IS, GMG, FS contributed to participant recruitment, clinical data management, safety assessment, bronchoscopy or other clinical assistance. ZX, FS secured the funding. MJ, ZX, FS wrote the manuscript.

## Acknowledgements

The work is supported by funds from the Canadian Institutes of Health Research (CIHR) Project & Foundation Programs and the Collaborative Health Research Program of CIHR and the Natural Sciences and Engineering Research Council of Canada. The authors are grateful to Tracy Tazzeo, Xueya Feng and Natallia Kazhdan for their technical assistance, and to all of the trial volunteers.

---

**Supplementary Figure 1.**
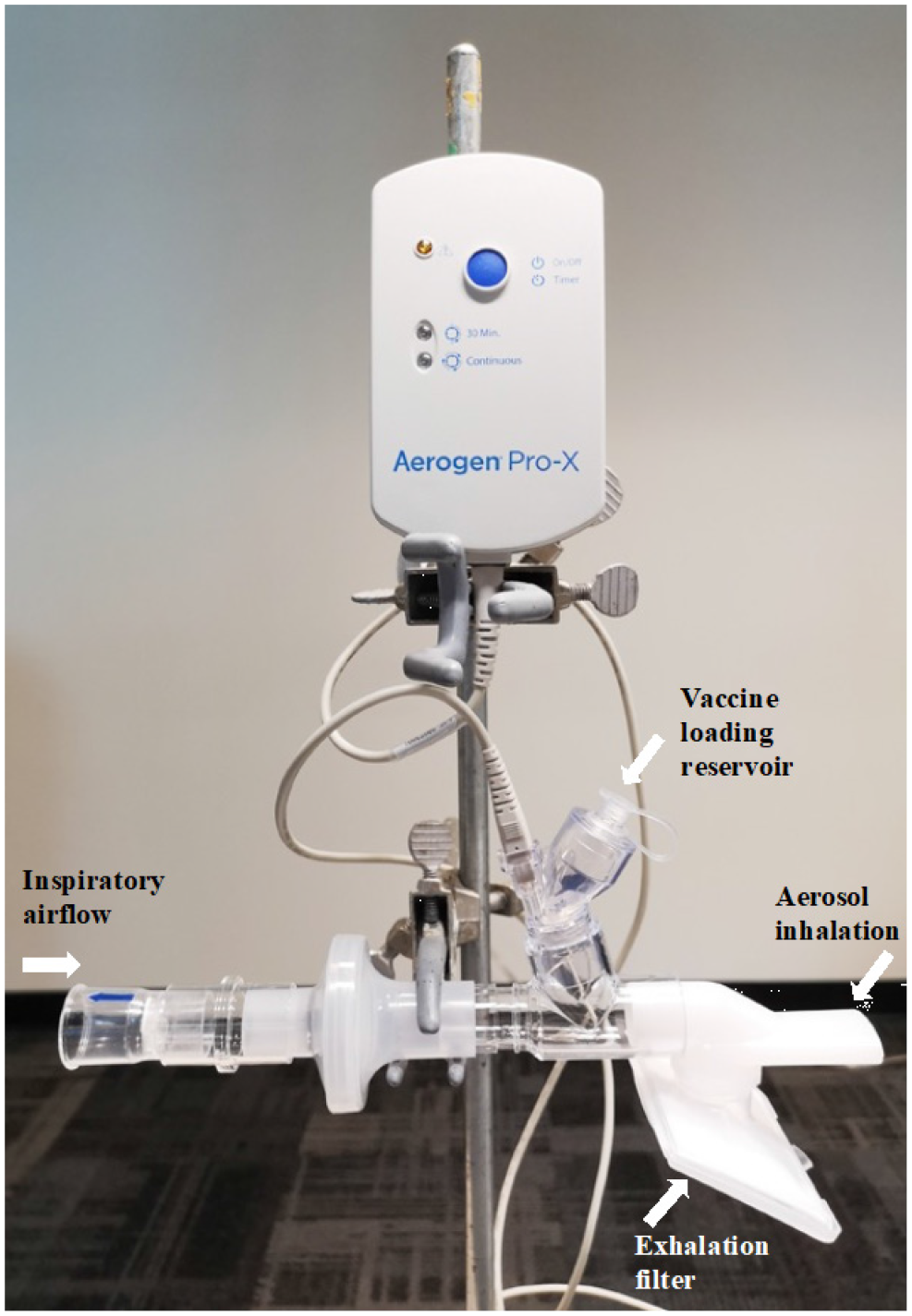
Inhaled aerosol device set-up. The Aeroneb® Solo Micropump was used as part of the delivery device set-up.

**Supplementary Figure 2.**
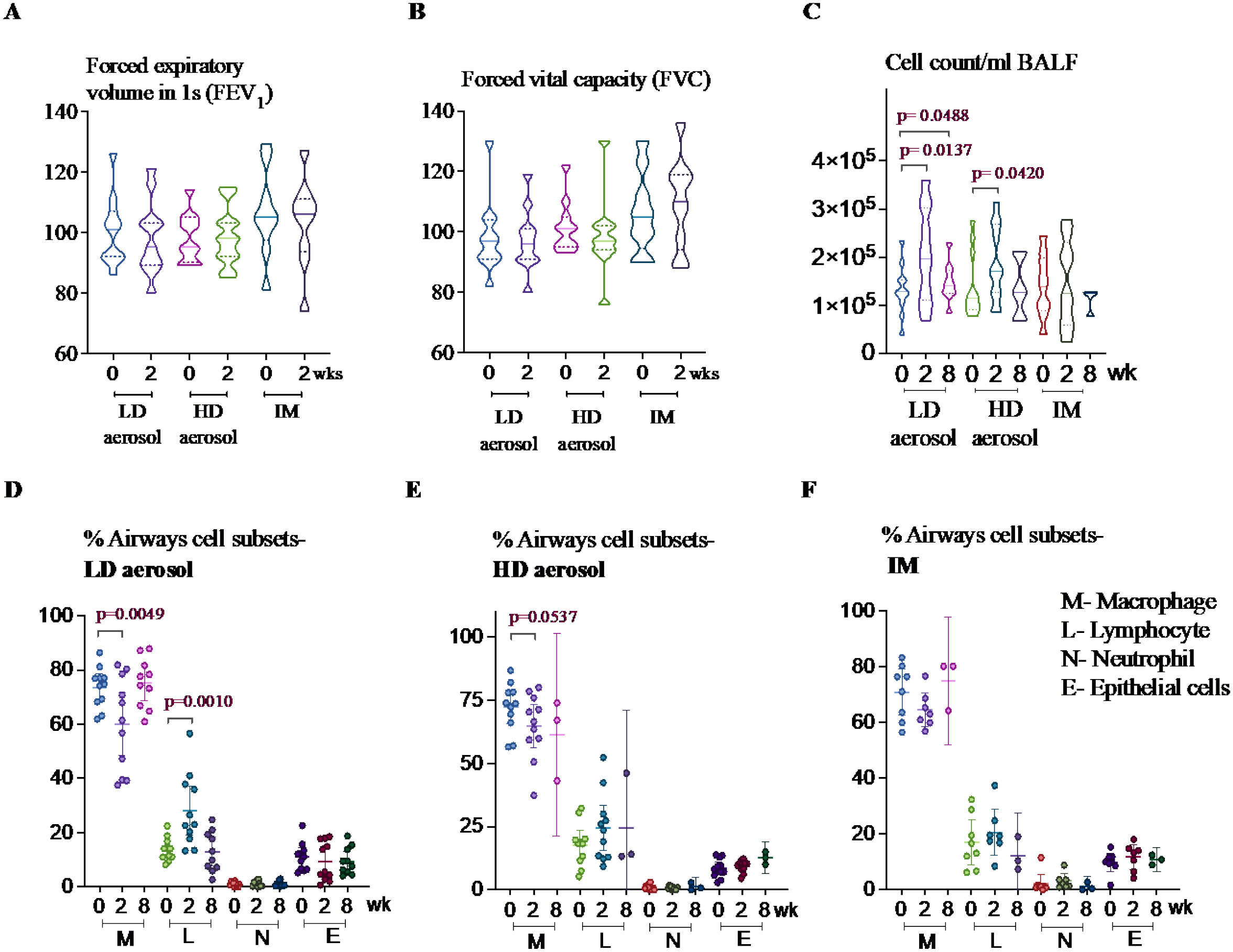
Respiratory function and bronchoalveolar cellular responses following aerosol or intramuscular vaccination. **A**), **B**) Lung function was assessed as FEV_1_ and FVC at baseline and 2wk post-LD aerosol, HD aerosol or IM vaccination. **C**), **D**), **F**) Frequencies of differential cells including macrophages, lymphocytes, neutrophils and epithelial cells in BALF from LD aerosol, HD aerosol and IM vaccine cohorts.

**Supplementary Figure 3.**
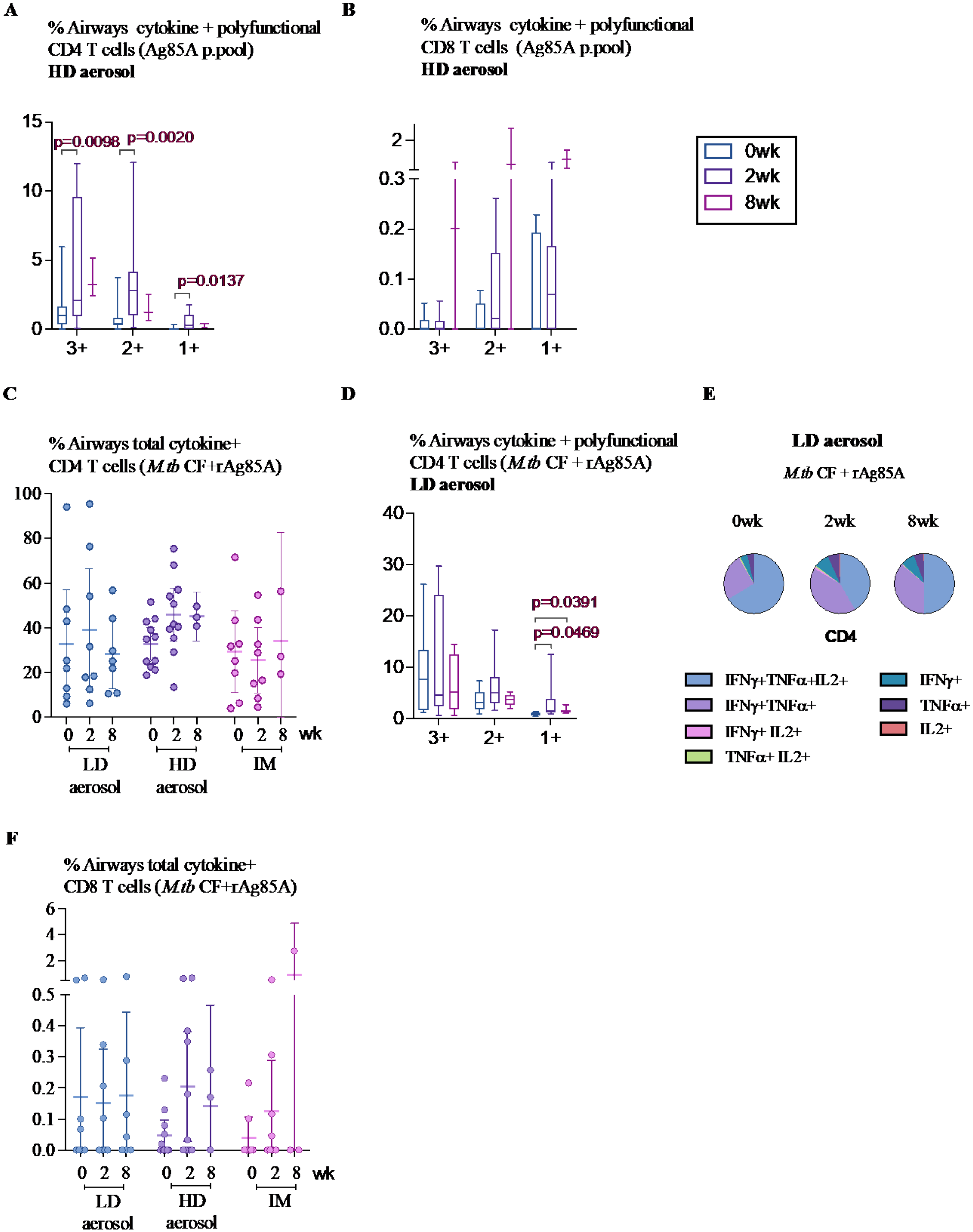
Induction of multifunctional T cells specific for Ag85A or a cocktail of mycobacterial antigens in the airways following aerosol or intramuscular vaccination. **A**), **B**) Frequencies of airways Ag85A-specific polyfunctional (triple/3+, double/2+ and single/1 cytokine-positive) CD4 and CD8 T cells at various timepoints in HD aerosol group. **C**) Frequencies of airways combined total cytokine-producing CD4 T cells specific for *M.tb* CF/rAg85A at various timepoints in LD aerosol, HD aerosol and IM cohorts. **D**) Frequencies of airways polyfunctional (triple/3+, double/2+ and single/1 cytokine-positive) CD4 T cells specific for *M.tb* CF/rAg85A at various timepoints in LD aerosol group. **E**) Median proportions displayed in pie chart of *M.tb* CF/rAg85A-specific airways CD4 T cells expressing a specific single or combination of two or three cytokines at various timepoints in LD aerosol group. **F**) Frequencies of airways combined total cytokine-producing CD8 T cells specific for *M.tb* CF/rAg85A at various timepoints in LD aerosol, HD aerosol and IM cohorts.

**Supplementary Figure 4.**
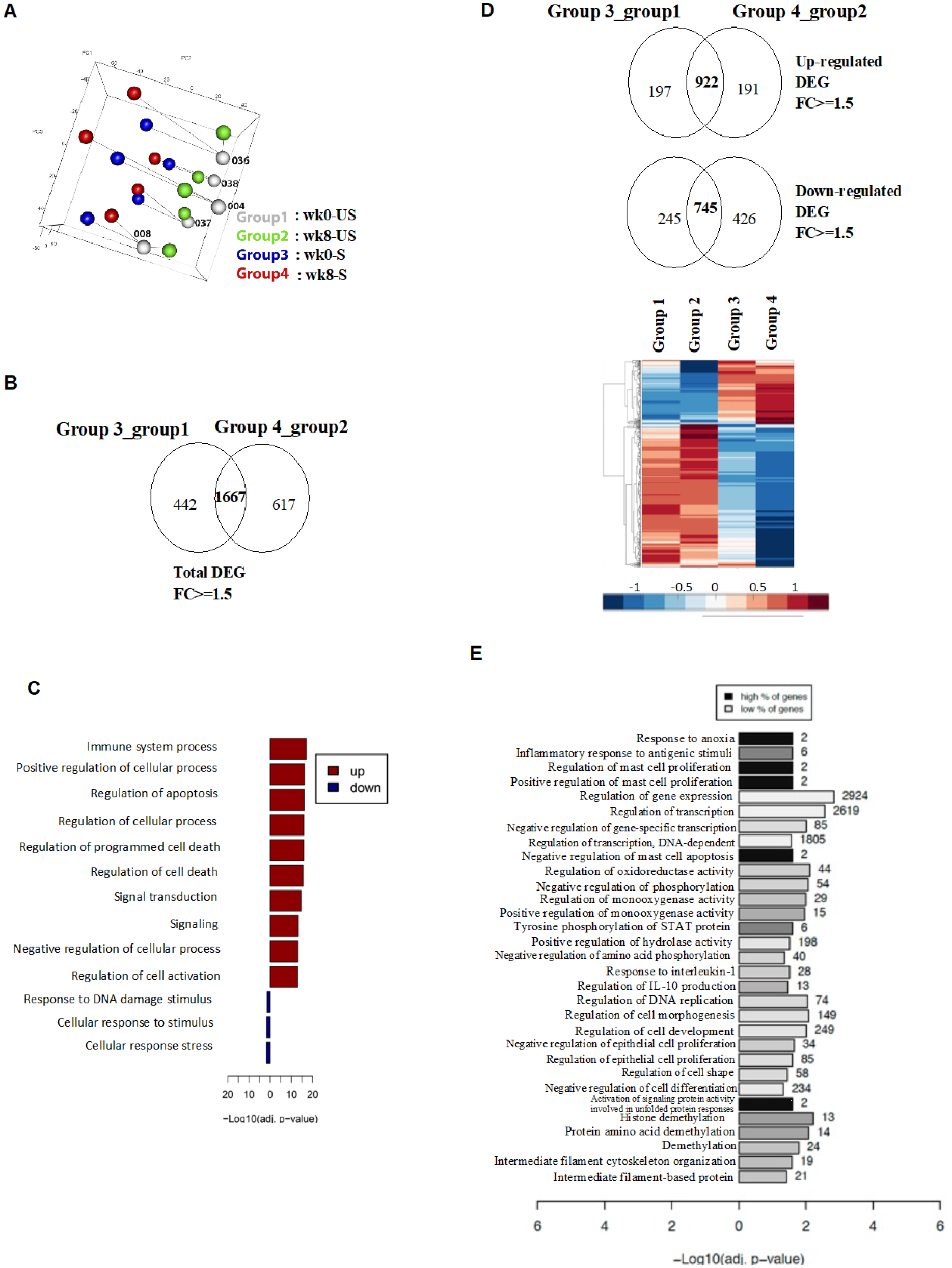
Transcriptomic analysis of alveolar macrophages (AM) following LD aerosol vaccination. **A**) Principal component analysis (PCA) of gene expression in AM obtained before (wk0) and after (wk8) LD aerosol vaccination cultured with (S) or without (US) stimulation. **B**) Venn diagram comparing all DEGs in pairwise comparison. **C**) Significantly enriched functional categories of biological processes by GO associated with DEGs shared between the baseline (wk0) Group 3/1 and aerosol vaccine (wk8) Group 4/2. **D**) Venn diagram comparing up- and down-regulated DEGs in pairwise comparison. Heatmap shows DEG uniquely up- and down-regulated, in stimulated aerosol (Group 4) AM. **E**) Significantly enriched functional categories of biological processes by GO associated with uniquely up-regulated DEGs in stimulated aerosol (Group 4) AM.

**Supplementary Figure 5.**
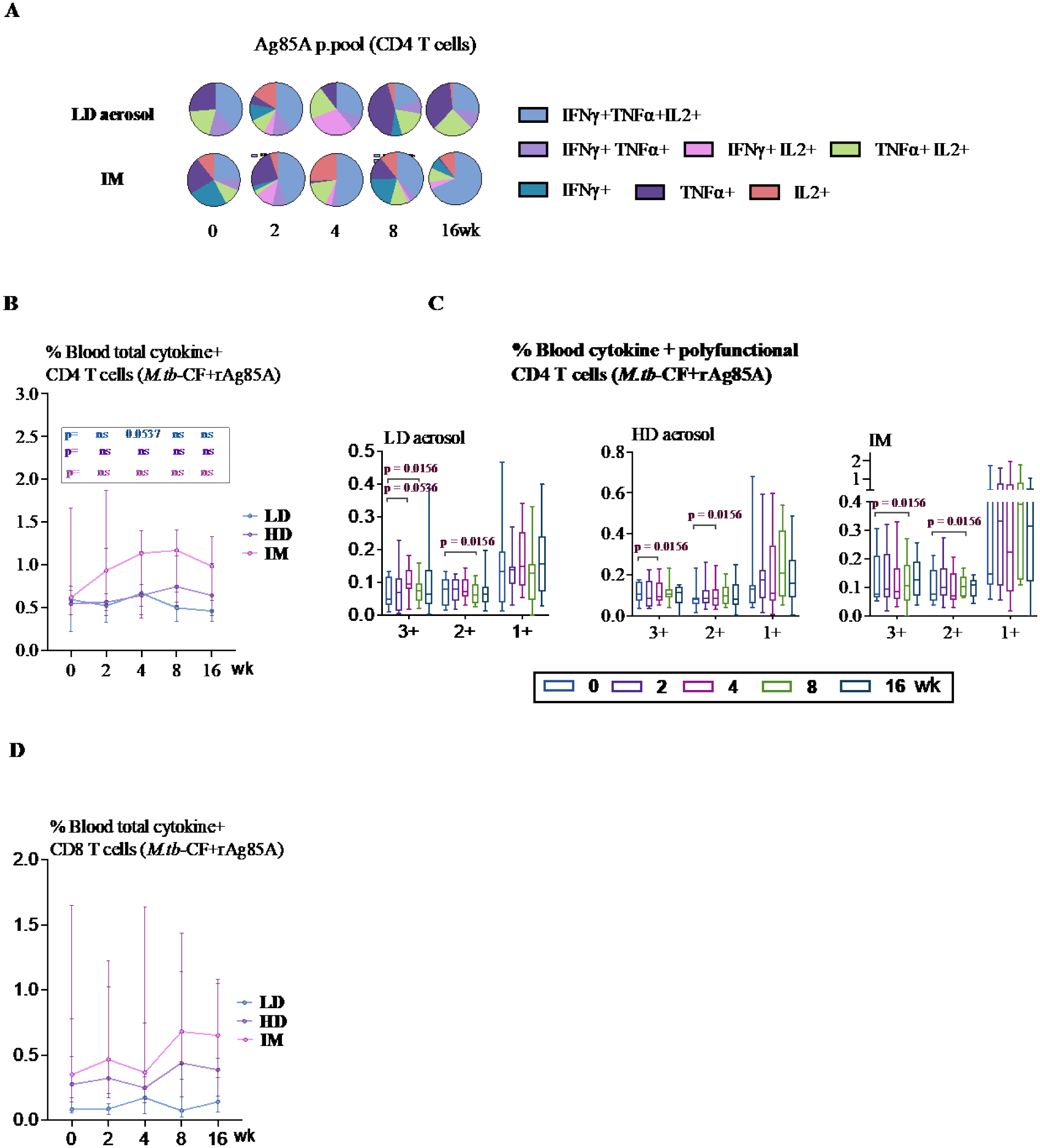
Induction of multifunctional T cells specific for Ag85A or a cocktail of mycobacterial antigens in the peripheral blood following aerosol or intramuscular vaccination. **A**) Median proportions displayed in pie chart of peripheral blood Ag85A-specific CD4 T cells expressing a specific single or combination of two or three cytokines at various timepoints in LD aerosol and IM vaccine groups. **B**) Frequencies of peripheral blood *M.tb* CF/rAg85A-specific combined total cytokine-producing CD4 T cells at various timepoints in LD aerosol, HD aerosol and IM cohorts. **C**) Frequencies of peripheral blood *M.tb* CF/rAg85A-specific polyfunctional (triple/3+, double/2+ and single/1 cytokine-positive) CD4 T cells at various timepoints in LD aerosol, HD aerosol and IM vaccine groups. **D**) Frequencies of peripheral blood *M.tb* CF/rAg85A-specific polyfunctional (triple/3+, double/2+ and single/1 cytokine-positive) CD8 T cells at various timepoints in LD aerosol, HD aerosol and IM vaccine groups.

**Supplementary Figure 6.**
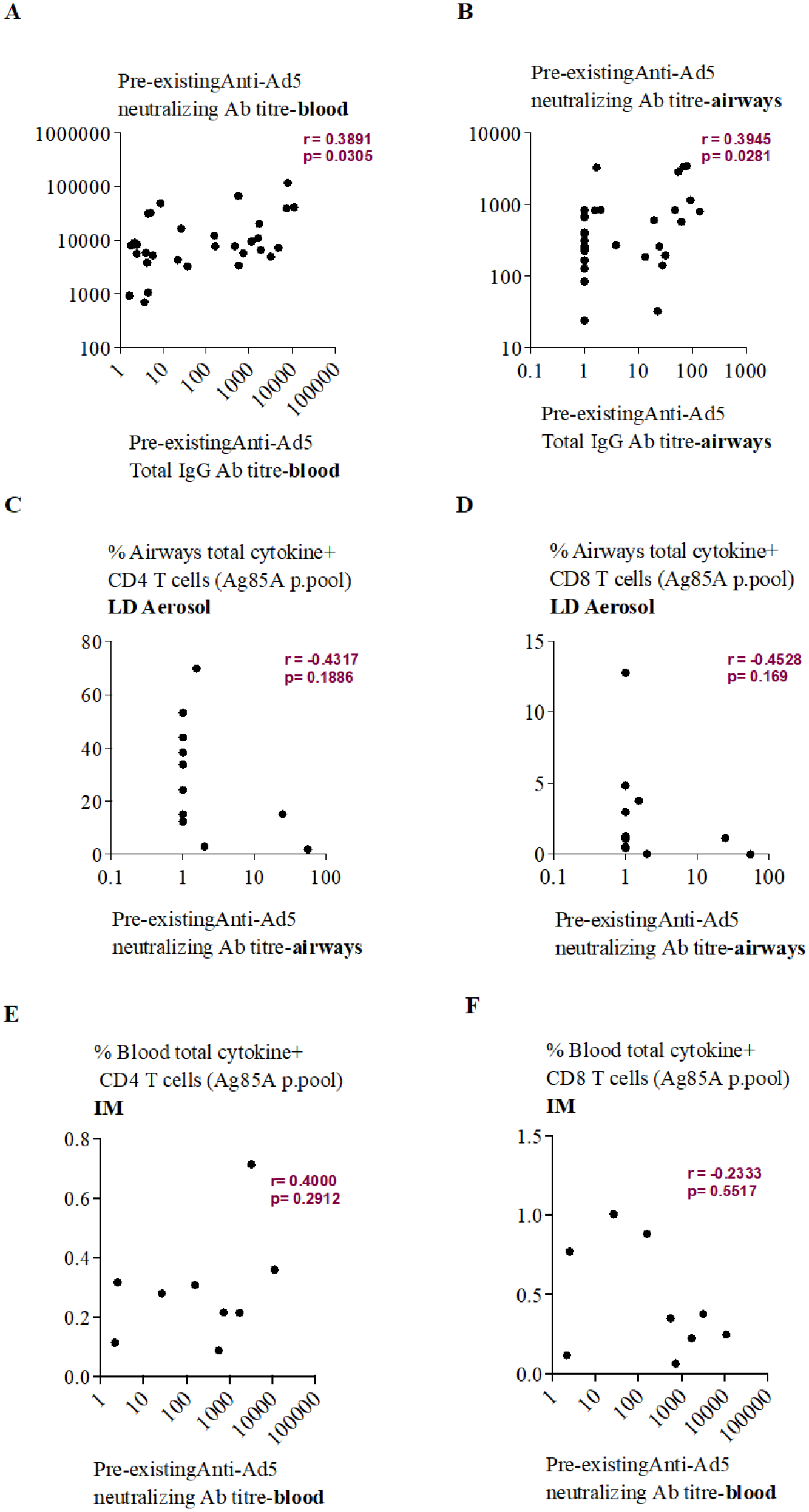
Impact of pre-existing anti-AdHu5 antibodies on the immunogenicity induced by LD aerosol and IM AdHu5Ag85A vaccination. **A**) Correlation plot of pre-existing anti-Ad5 neutralizing antibodies versus pre-existing anti-Ad5 total IgG antibody titers in the blood of all trial participants. **B**) Correlation plot of pre-existing anti-Ad5 neutralizing antibodies versus pre-existing anti-Ad5 total IgG antibody titers in the airways of all trial participants. **C**) Correlation plot of pre-existing anti-Ad5 neutralizing antibodies versus peak airways antigen-specific total cytokine+ CD4 T cell responses in LD aerosol vaccine group. **D**) Correlation plot of pre-existing anti-Ad5 neutralizing antibodies versus peak airways antigen-specific total cytokine+ CD8 T cell responses in LD aerosol vaccine group. **E**) Correlation plot of pre-existing anti-Ad5 neutralizing antibodies versus peak peripheral blood antigen-specific total cytokine+ CD4 T cell responses in IM vaccine group. **F**) Correlation plot of pre-existing anti-Ad5 neutralizing antibodies versus peak peripheral blood antigen-specific total cytokine+ CD8 T cell responses in IM vaccine group.

